# Higher COVID-19 vaccination rates are linked to decreased county-level COVID-19 incidence across USA

**DOI:** 10.1101/2021.03.05.21252946

**Authors:** Arjun Puranik, AJ Venkatakrishnan, Colin Pawlowski, Bharathwaj Raghunathan, Eshwan Ramudu, Patrick Lenehan, Vineet Agarwal, Savita Jayaram, Mayank Choudhary, Venky Soundararajan

## Abstract

Real world evidence studies of mass vaccination across health systems have reaffirmed the safety^1^ and efficacy^2,3^ of the FDA-authorized mRNA vaccines for COVID-19. However, the impact of vaccination on community transmission remains to be characterized. Here, we compare the cumulative county-level vaccination rates with the corresponding COVID-19 incidence rates among 87 million individuals from 580 counties in the United States, including 12 million individuals who have received at least one vaccine dose. We find that cumulative county-level vaccination rate through March 1, 2021 is significantly associated with a concomitant decline in COVID-19 incidence (Spearman correlation ρ = −0.22, p-value = 8.3e-8), with stronger negative correlations in the Midwestern counties (ρ = −0.37, p-value = 1.3e-7) and Southern counties (ρ = −0.33, p-value = 4.5e-5) studied. Additionally, all examined US regions demonstrate significant negative correlations between cumulative COVID-19 incidence rate prior to the vaccine rollout and the decline in the COVID-19 incidence rate between December 1, 2020 and March 1, 2021, with the US western region being particularly striking (ρ = −0.66, p-value = 5.3e-37). However, the cumulative vaccination rate and cumulative incidence rate are noted to be statistically independent variables, emphasizing the need to continue the ongoing vaccination roll out at scale. Given confounders such as different coronavirus restrictions and mask mandates, varying population densities, and distinct levels of diagnostic testing and vaccine availabilities across US counties, we are advancing a public health resource to amplify transparency in vaccine efficacy monitoring (https://public.nferx.com/covid-monitor-lab/vaccinationcheck). Application of this resource highlights outliers like Dimmit county (Texas), where infection rates have increased significantly despite higher vaccination rates, ostensibly owing to amplified travel as a “vaccination hub”; as well as Henry county (Ohio) which encountered shipping delays leading to postponement of the vaccine clinics. This study underscores the importance of tying the ongoing vaccine rollout to a real-time monitor of spatio-temporal vaccine efficacy to help turn the tide of the COVID-19 pandemic.

---

As mass vaccination efforts ramp up worldwide to combat the COVID-19 pandemic, there is a need for real-time assessment of how factors such as vaccination rates impact SARS-CoV-2 community transmission and localized outbreaks. As of March 2, 2021, over 51 million individuals in the United States have received at least one COVID-19 vaccine dose, including over 26 million individuals who have been fully vaccinated with both doses of the Moderna (mRNA-1273) or Pfizer/BioNTech (BNT162b2) vaccines^4^. While these mRNA vaccines were shown to decrease the risk for symptomatic COVID-19 in Phase 3 clinical trials^5,6^, we and others have recently reaffirmed their real world efficacy and safety via holistic inference from context-rich electronic health records (EHRs) across American and Israeli health systems^2,7^.

In order to conduct a real-world analysis of the impact of vaccination on COVID-19 incidence in the USA, we first obtained county-level PCR testing data from the *CDC COVID Data Tracker*^*4*^. We also obtained temporal and cumulative county-level vaccination data (number of patients who have received at least 1 vaccine dose) from *CovidActNow*^*8*^ as of March 1, 2021. The *CovidActNow* dataset includes vaccine administration data from 580 counties from the District of Columbia and 13 US states including: Georgia, Maine, Maryland, Minnesota, Missouri, Nevada, New Jersey, New York, Ohio, Oregon, Pennsylvania, Washington, and Wyoming, with a total population of over 87 million. This includes over 12 million individuals who have received at least one dose of either the Moderna or Pfizer/BioNTech vaccines.

Contrasting the 7-day PCR positivity rates in the week leading up to March 1, 2021 (current vaccination phase) vs. the week leading up to December 1, 2020 (pre-vaccination phase), we observe a sharp decline in COVID-19 incidence across the country (**Figure 1A**). Numerous factors that are unrelated to the vaccine rollout may have contributed to this decline, hence necessitating quantification of vaccination impact on this trend. In order to assess the impact of the vaccination on localized COVID-19 incidence, we examined the correlation between two metrics: (i) *Cumulative county-level vaccination rate*: percentage of the county population who have received at least one dose, and (ii) *December-March change in COVID-19 incidence rate*: Difference in 7-day COVID-19 incidence rates from December 1, 2020 to March 1, 2021. The 7-day COVID-19 incidence rate is the number of reported COVID-19 cases in the county per 10,000 individuals averaged over the past 7 days. We find that the cumulative county-level vaccination rate is significantly negatively correlated with the December-March change in COVID-19 incidence rate (**Figure 1B;** Spearman’s ρ = −0.22, p-value: 8.3e-08; **Supplementary Figure S1** − with 95% confidence intervals added to the y-axis). We also independently performed a manual curation of cumulative vaccination rates through March 1, 2021 across 1408 counties from 28 states, and the negative correlation with the December-March change in COVID-19 incidence rate was consistent (**Supplementary Figure S2**). This shows that higher COVID-19 vaccination rates are associated with decreased county-level COVID-19 incidence for these counties in the United States.

**Figure 1.**
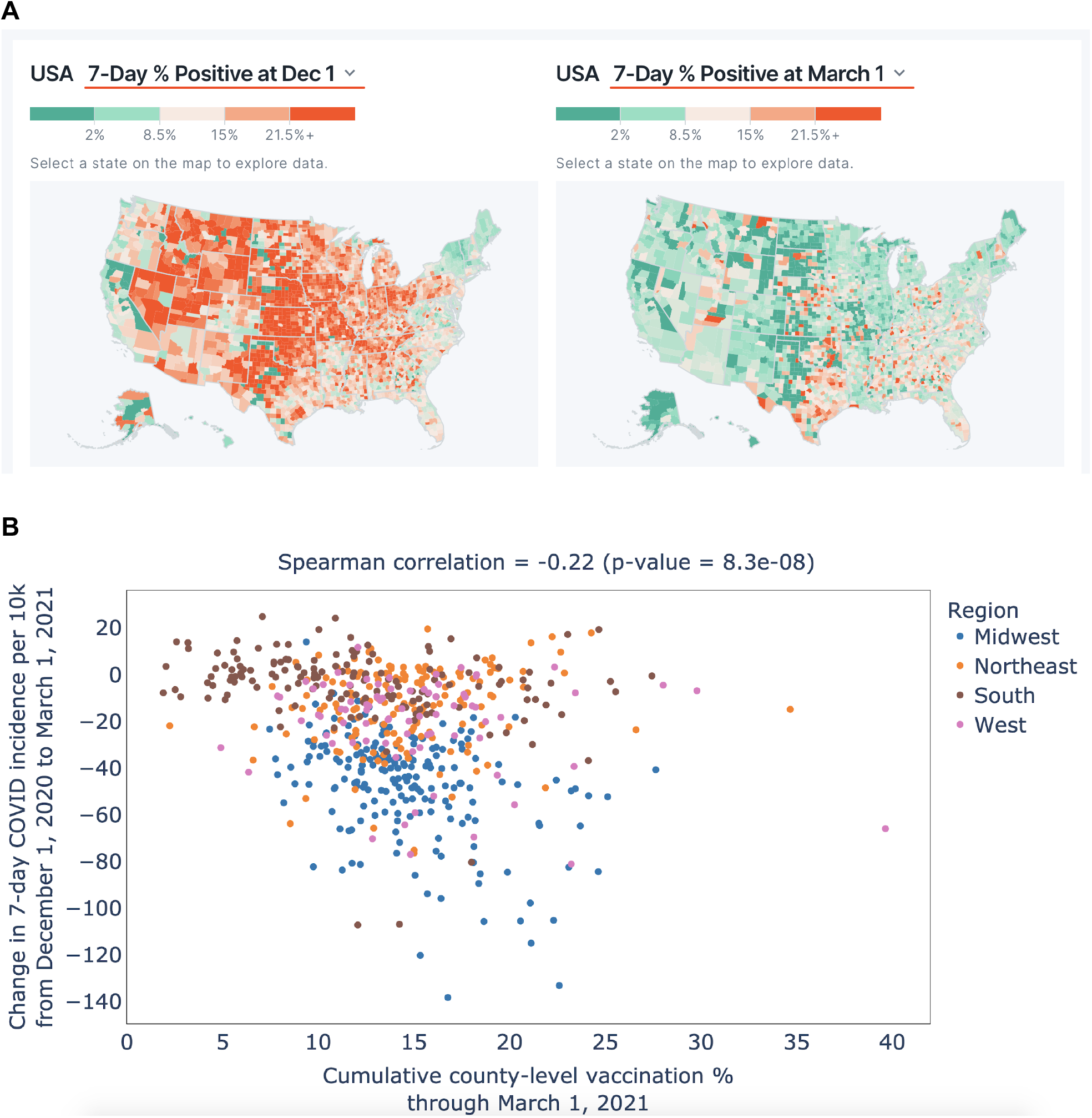
Relationship between mass vaccination and COVID-19 incidence. **(A)** US map of counties showing 7-day SARS-CoV-2 PCR positivity rates (PPR) for December 1 2020 prior to onset of FDA-authorized COVID-19 vaccine rollout (left panel), and 7-day PPR for March 1 2021 after Phase 1a of mRNA vaccination between December 2020 and February 2021. The colorbar varies from *green* (low PPR) to *red* (high PPR). **(B)** Scatterplot shows the relationship between cumulative vaccination rate as of March 1, 2021 and difference between the 7-day COVID incidence (per 10,000 individuals) between December 1, 2020 and March 1, 2021. This includes 580 counties (cumulative population: 88 million) from 14 states. Counties are colored according to the US census bureau region^9^ of their respective states. The *nferX vaccination monitor* resource to create the above visuals is available for free from https://public.nferx.com/covid-monitor-lab/vaccinationcheck.

To evaluate the impact of vaccination in different parts of the country, we grouped the counties into four regions^9^: (i) Midwest: Ohio, Minnesota, Missouri; (ii) South: District of Columbia, Georgia, Maryland; (iii) West: Nevada, Oregon, Washington, Wyoming; and (iv) Northeast: Maine, New Jersey, New York, Pennsylvania. Interestingly, there is a significant negative correlation between the cumulative vaccination rate and the December-March change in COVID-19 incidence rate for the Midwest (**Figure 2A**; ρ=−0.37, p-value: 1.3e-7) and the South (**Figure 2B;** ρ=−0.33, p-value: 4.5e-5). The counties in the West (**Figure 2C;** ρ =−0.06, p-value: 0.61) and Northeast (**Figure 2D;** ρ=0.19, p-value: 0.2) regions do not demonstrate any substantial correlation.

**Figure 2:**
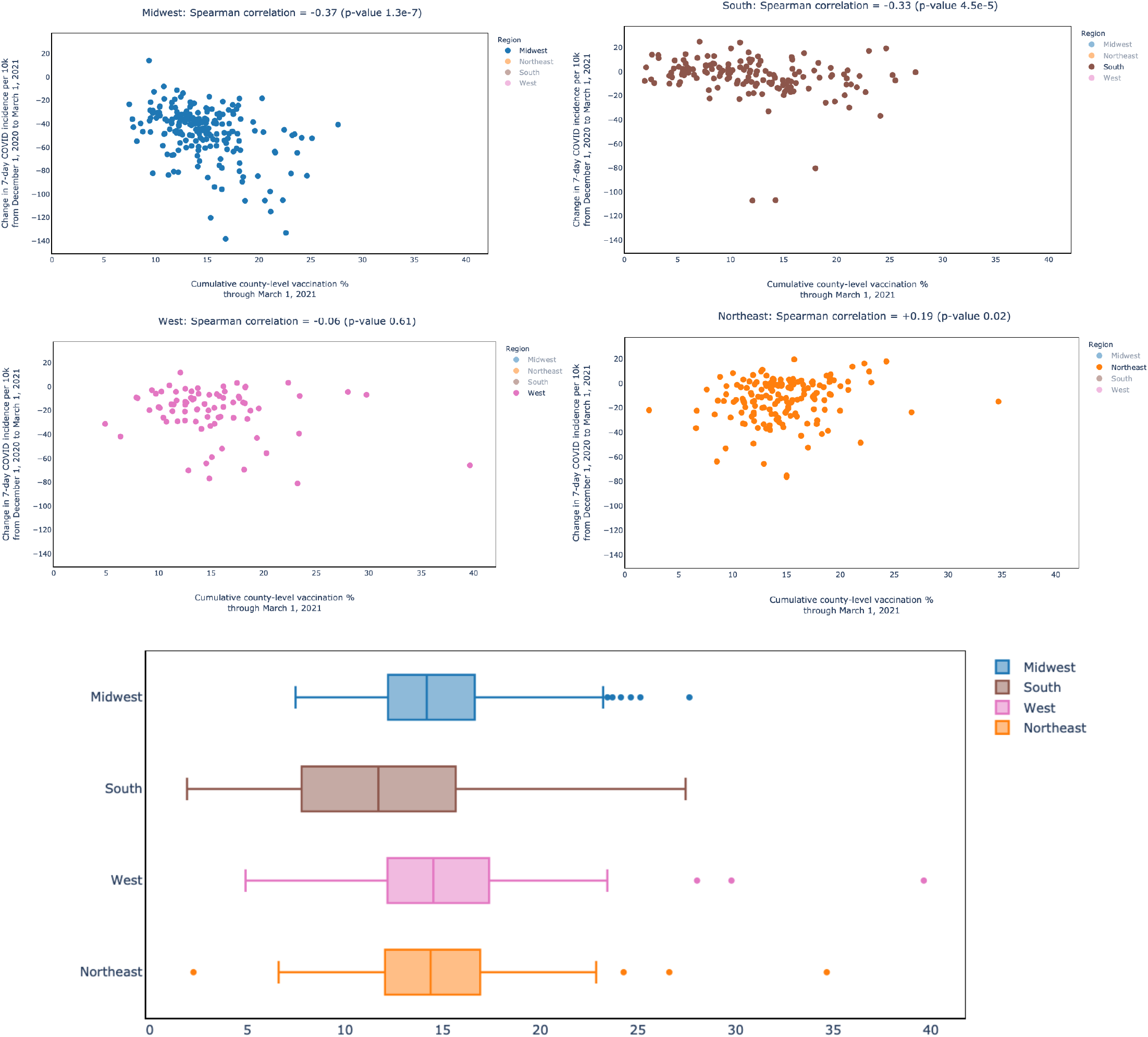
Cumulative vaccination through March 1, 2021 versus change in incidence from December 1, 2020 to March 1, 2021, broken down by region. Shown are region-specific scatter plots & Spearman correlation. These are sorted in order of low to high Spearman correlation: **(A) Midwest**: Minnesota, Missouri, Ohio (194 counties present); **(B) South**: District of Columbia, Georgia, Maryland (150 counties present); **(C) West**: Nevada, Oregon, Washington, Wyoming (79 counties present); **(D) Northeast**: Maine, New Jersey, New York, Pennsylvania (157 counties present). **(E)** Cumulative vaccination percentages through March 1, 2021 for the 4 regions as boxplots. The *nferX vaccination monitor* resource to create the above visuals is available for free from https://public.nferx.com/covid-monitor-lab/vaccinationcheck.

Given that the above analysis does not factor in pre-existing immunity from prior exposure to SARS-CoV-2, and particularly since certain parts of the country experienced more severe outbreaks during earlier phases of the pandemic (e.g., New York, Seattle), we next looked at the relationship between cumulative county-level incidence rate up to December 1, 2020 and the change in COVID-19 incidence rates (December 1, 2020 to March 1, 2021). While we find that there is a significant negative correlation in the counties across all the four regions, the US western region counties are particularly distinctive in their significance (**Figure 3A-E**). Although cumulative COVID-19 incidence through prior infection waves appears to be associated with the ongoing decline of county-level COVID-19 incidence, it must be noted that this phenomenon is independent of the cumulative vaccination rate (**Supplementary Figure S3**). While differential levels of prior COVID infectivity levels and ongoing vaccine rollout may consequently both factor into the distinct county-level trends in SARS-CoV-2 positivity, their statistical independence emphasizes the need to continue mass vaccination at scale.

**Figure 3.**
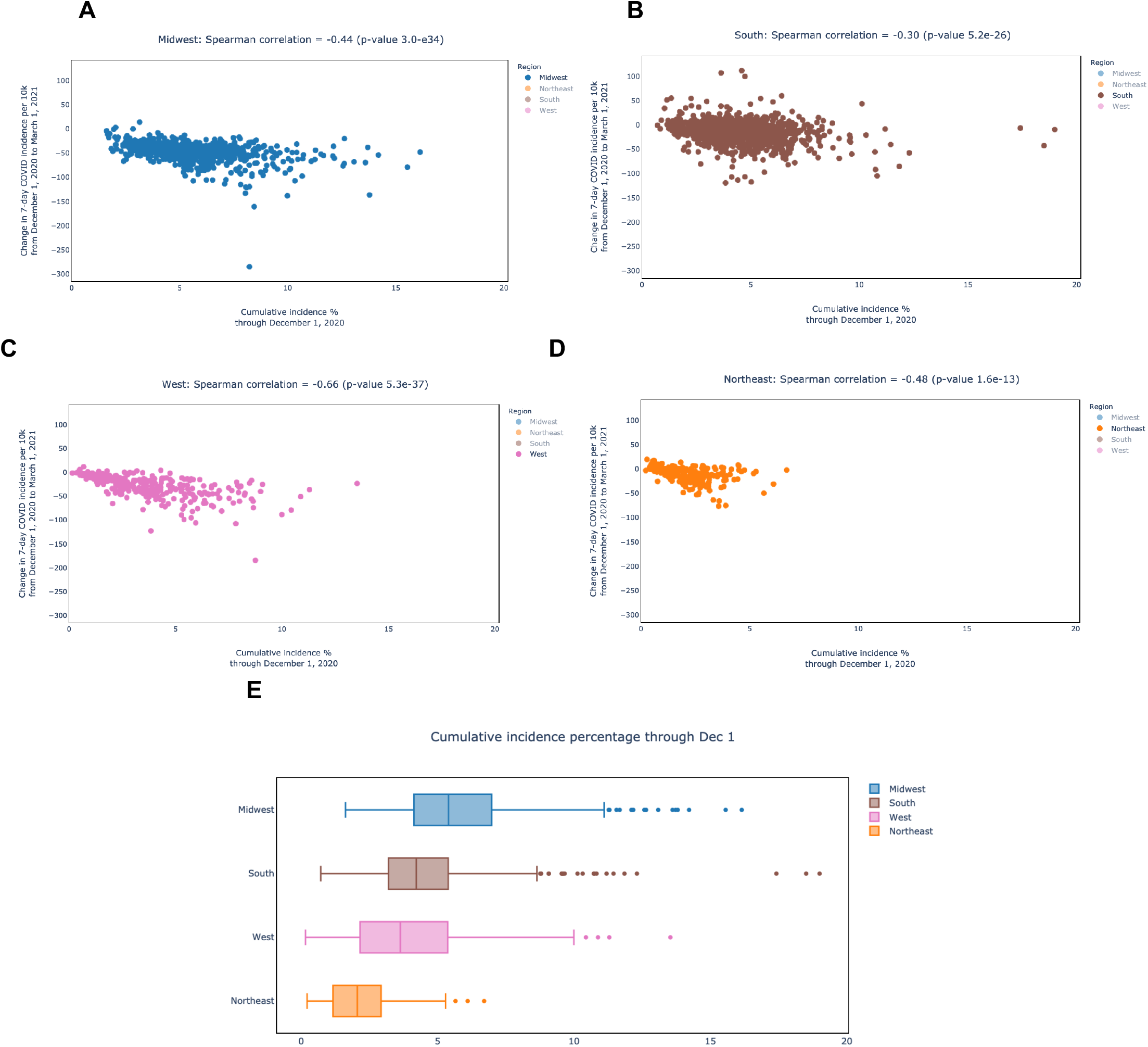
Cumulative county-level COVID-19 incidence through December 2020 (pre-vaccination phase) as a function of the recent change in incidence from December 1, 2020 to March 1, 2021 (phase 1a vaccination period). Scatter plots are shown for US counties in the following regions: **(A)** Midwest, **(B)** South, **(C)** West, and **(D)** Northeast. **(E)** Boxplots of cumulative COVID-19 incidence rate through December 1, 2020 for US counties in these four regions. The *nferX vaccination monitor* resource to create the above visuals is available for free from https://public.nferx.com/covid-monitor-lab/vaccinationcheck.

In an attempt to account for the time taken for effect of vaccination on the decline in the prevailing county level incidence rates (as of March 1, 2021), we next allowed a one-month “gap period”, analyzing the effect of cumulative COVID-19 vaccination until only February 1, 2021. This temporal record of COVID-19 vaccination as of February 1, 2021 is available at the county level from Texas, Ohio, Michigan, North Carolina, and District of Columbia. Analysis of this data shows that the cumulative county-level vaccination rate until February 1, 2021 is significantly negatively correlated with the December 1, 2020 to March 1, 2021 change in COVID-19 incidence rate (**Supplementary Figure S4;** ρ =−0.27, p-value=3.5e-8). This preliminary analysis at the early phases of vaccine rollout incorporating a time-delay for vaccine-induced immunity to manifest does underline the positive impact of vaccine administration on the decline of COVID-19 incidence. The 1-month “gap period” used for this analysis is purely exemplary and may require adjusting for various factors such as distinct mechanisms of the 2-dose mRNA vaccines from Moderna and Pfizer/BioNTech versus, for example, the recently FDA-authorized 1-dose vaccine from Johnson & Johnson (J&J)^10^.

We note that there are a number of extrinsic factors that could confound the associations observed in this study. For example, county-level policies on social distancing norms and their enforcement could impact COVID-19 incidence^11^. Rates of PCR-testing and case reporting are important confounders as well, which are analyzed through a preliminary analysis of the available percent positivity rates (PPR) at the county-level (**Supplementary Figure S5**). The COVID-19 cases that were asymptomatic and undiagnosed clinically cannot be accounted for in this study. There is variation in terms of demographic factors (e.g., age above 65) and population density between counties. Socio-economic differences between counties such as level of access to healthcare and insurance could also affect the findings presented here. In addition, study design choices such as the time period for the analysis (December 1, 2020 to March 1, 2020) could impact the findings as well. To address this issue, we conducted a sensitivity analysis varying the start date for the study period, including November 15, 2020, December 1, 2020 (actual study start date selected), December 15, 2020, January 1, 2021, January 15, 2021 (**Supplementary Figure S6**). We observe that the correlation between cumulative vaccination rate and change in 7-day COVID-19 incidence changes for each of the four regions as the study start date varies, which may be explained by different vaccine rollout times in these regions. An analysis that takes into account particular vaccine rollout dates for different counties could help us better understand these distinct spatio-temporal trends.

Given the evolving situation with COVID-19 incidence and vaccination rates, there is an imminent need to monitor the real-time impact at a county level in order to guide public health measures and policy decisions. Towards this, we are advancing a public health resource for amplifying transparency in vaccine efficacy monitoring tied to spatio-temporal news feeds (https://public.nferx.com/covid-monitor-lab/vaccinationcheck). Applying this resource to analyze outliers in our vaccine efficacy analysis highlights examples such as Dimmit County in Texas (**Figure 4A**) and Henry County in Ohio (**Figure 4B**). In Dimmit County, there was a surprising increase in COVID-19 incidence between December-March while a majority of the other Texas counties underwent significant decreases in incidence, despite Dimmit being one of the counties with highest vaccination rates in Texas. The associated news feed on our app^1^ suggests that, by virtue of being a “vaccine hub”, extensive out-of-county travel was associated with this county during the CDC-mandated Phase-1a vaccination period (**Figure 4A**). Another interesting case study from use of our resource to study outliers is Henry County in Ohio, that displays a significant increase in COVID-positivity from pre-vaccination (December 1, 2020) to current time (March 1, 2021), despite its neighboring counties significantly improving their COVID-positivity during that time period. The associated news feed on the app^2^ suggests that the COVID vaccine clinics in Henry county were postponed owing to shipment delays, which may explain in part the disturbing trend vis-a-vis the prevailing COVID incidence in this county (**Figure 4B**). Finally, an exemplar of a highly vaccinated neighborhood is Churchill County in Nevada (**Figure 4C**), which displays a concomitant decrease in COVID incidence during the Phase 1a vaccination period^3^.

**Figure 4.**
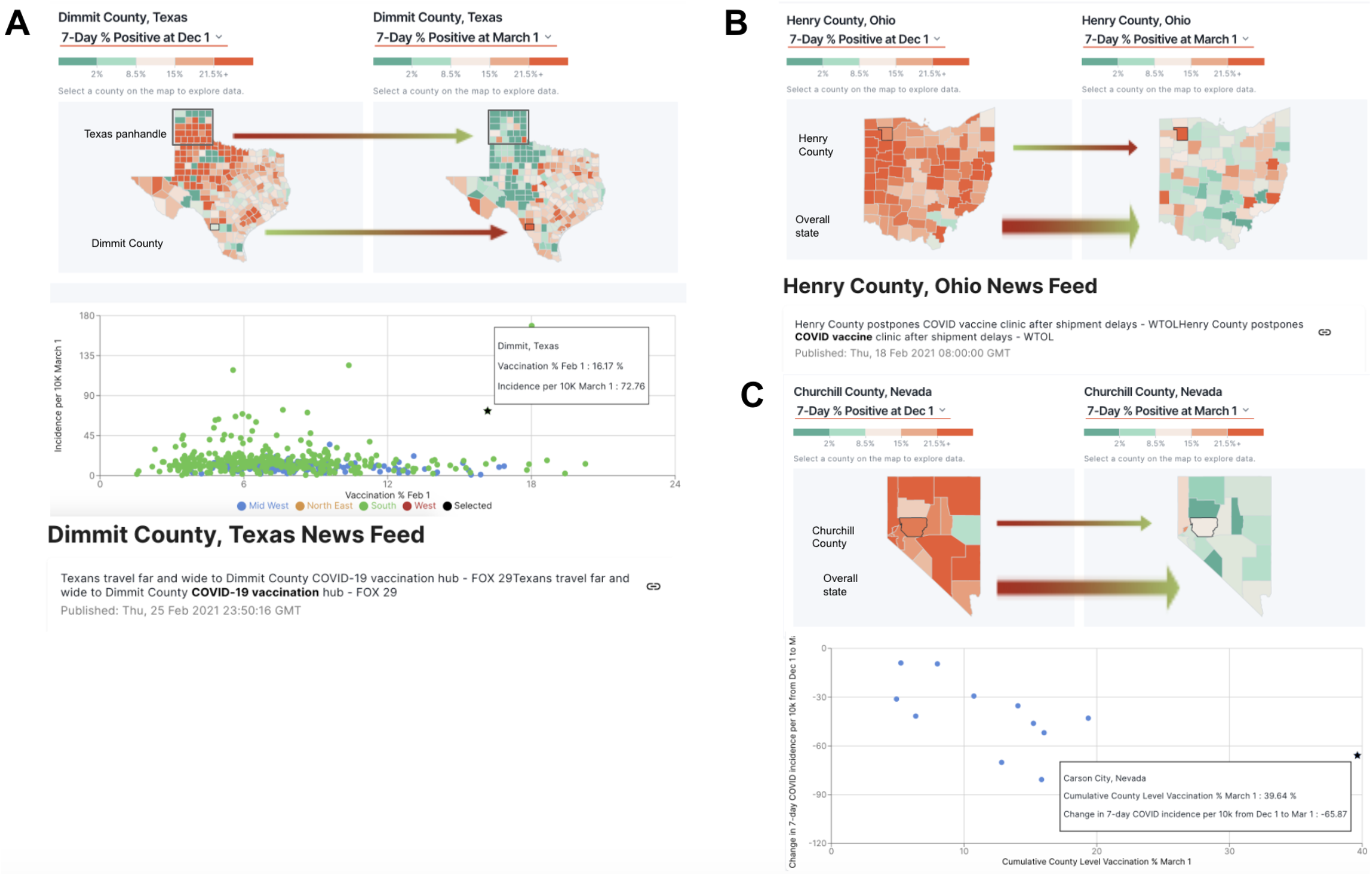
nferX Vaccinations Monitor app showing cumulative vaccination rates, PCR positivity rates, and personalized news feeds for select US counties. In the maps, counties are colored by their 7-day PCR percentage positivity rates (PPR) on December 1, 2020 and March 1, 2021. In the scatterplot of the counties, the cumulative vaccination percentage through February 1, 2021 is plotted against the incidence of COVID-19 per 10,000 individuals on March 1, 2021. The spatiotemporally personalized news feeds show recent articles about the selected counties. **(A) Dimmit County, Texas:** In the upper frame, we show how the 7-day PPR has largely declined for the counties in the Texas panhandle, but increased for Dimmit County, Texas. In the middle frame, we observe that Dimmit County has a relatively high vaccination rate (16.2% as of February 1, 2021) and incidence rate (72.8 cases per 10K as of March 1, 2021). In the lower frame, we show recent news articles which indicate that this county is a vaccination hub. **(B) Henry County, Ohio:** In the upper frame, we show how the 7-day PPR has largely declined for the counties in Ohio, but increased for Henry County, Ohio. In the lower frame, we show recent news articles which indicate that this county has experienced COVID-19 vaccine shipment delays. **(C) Churchill County, Nevada:** In the upper frame, we show how the 7-day PPR has largely declined for counties in the state of Nevada, including Churchill County. In the lower frame, we show a scatterplot of the cumulative county-level vaccination percentage as of March 1, 2021 vs. the change in 7-day COVID-19 incidence rate per 10K individuals from December 1, 2020 to March 1, 2021. The nferX vaccination monitor resource to create the above visualizations is available for free from the following website −- https://public.nferx.com/covid-monitor-lab/vaccinationcheck.

Even as people await their turn to get vaccinated, there are reports of subpopulations declining to get vaccinated owing misinformation around the vaccines’ safety and efficacy, which is being amplified through social media^12^. On the heels of the real-world based evidence on the vaccine’s safety^1^ and the efficacy^2,3^ for the vaccinated individuals, this study and the associated resource we have built emphasizes the positive public health impact of the vaccination campaign and further motivates the rapid vaccine rollout globally to quell the COVID-19 pandemic comprehensively.

## Data Availability

After publication, the data will be made available to others upon reasonable requests to the corresponding author. A proposal with a detailed description of study objectives and the statistical analysis plan will be needed for evaluation of the reasonability of requests.

## Supplementary Material

**Supplementary Figure S1.**
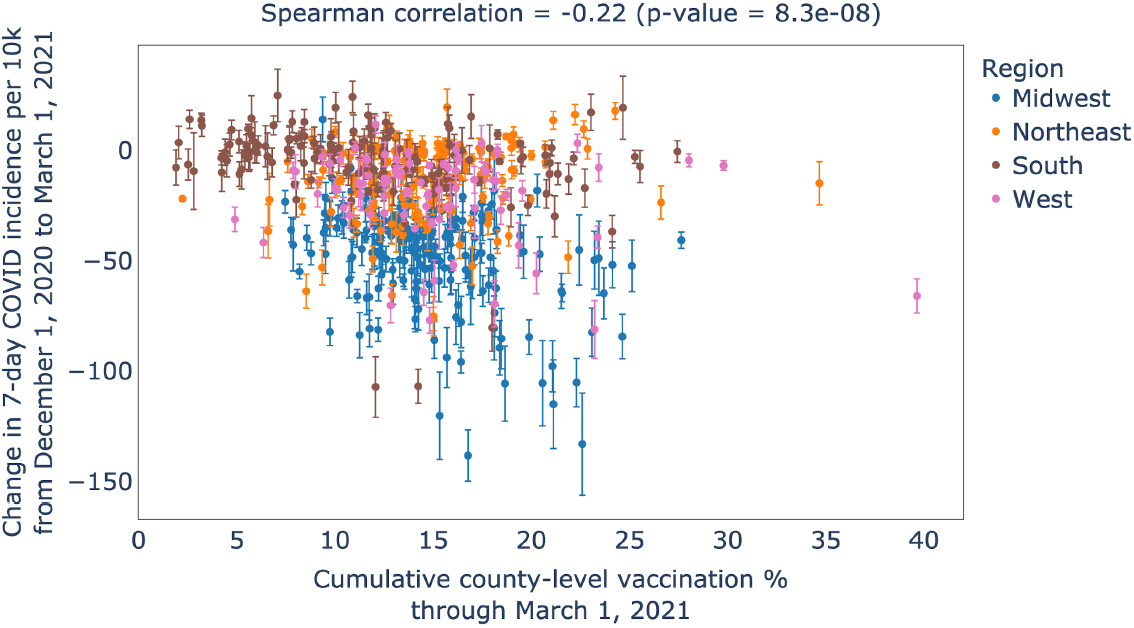
Cumulative county-level vaccination through March 1 2021 versus the change in 7-day COVID-19 incidence between December 1, 2020 and March 2021. This analysis included 580 counties from the District of Columbia and 13 states. The 95% confidence intervals to the y-axis (change in 7-day COVID incidence per 100,000 individuals) are shown.

**Supplementary Figure S2.**
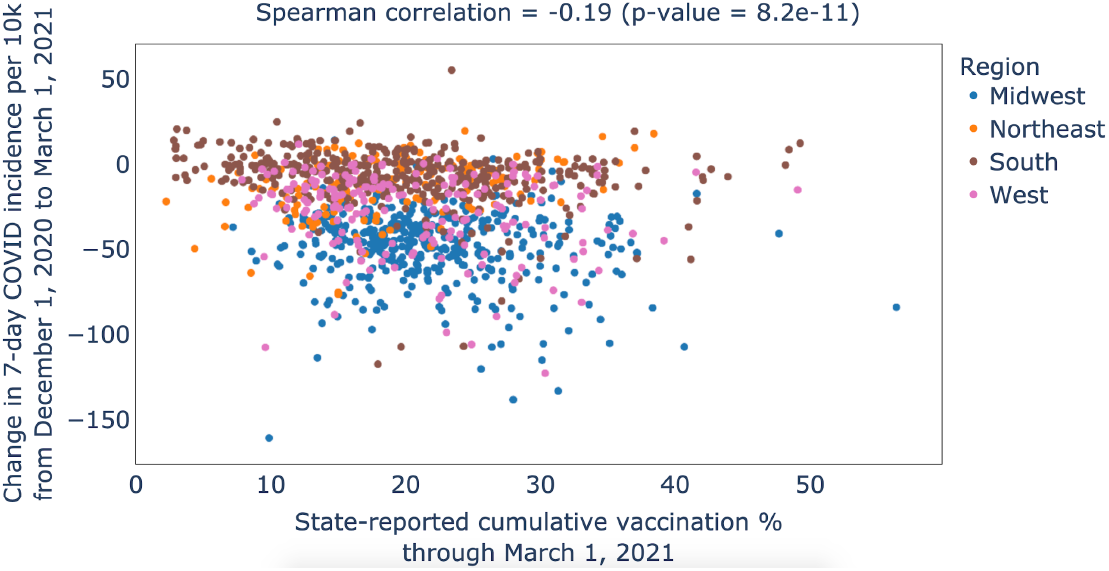
Cumulative county-level vaccination through March 1 2021 versus the change in 7-day COVID-19 incidence between December 1, 2020 and March 2021. This analysis includes 1987 counties from 40 states.

**Supplementary Figure S3.**
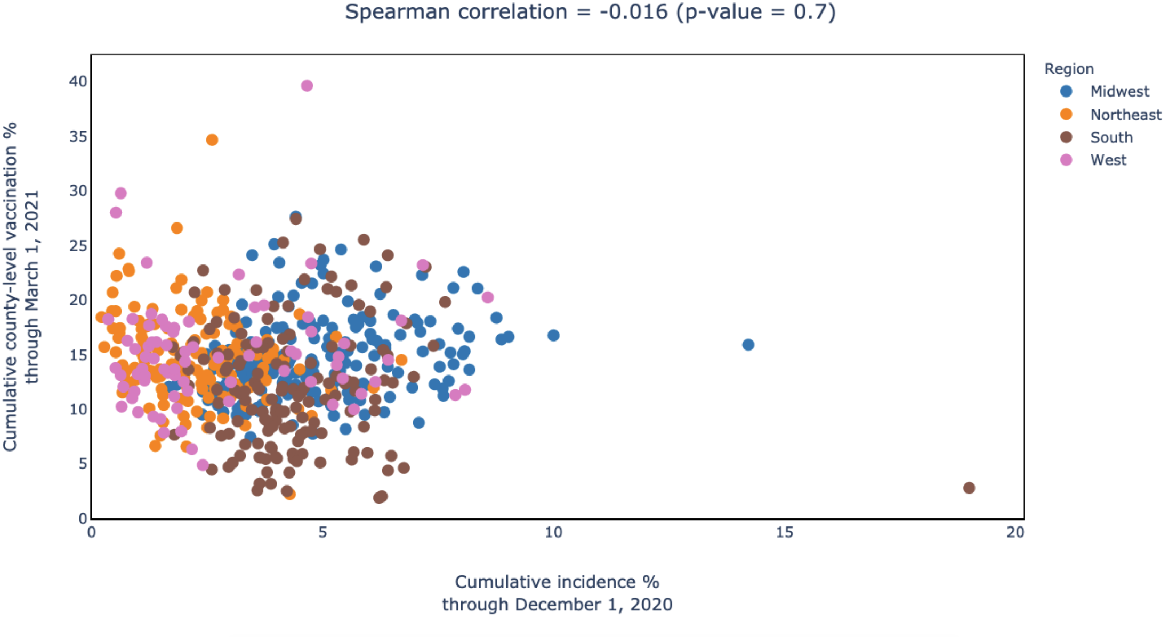
Cumulative COVID-19 incidence rate through December 1, 2020 versus cumulative vaccination percentage through March 1, 2021. The lack of correlation between these two factors emphasizes the independence of these variables.

**Supplementary Figure S4.**
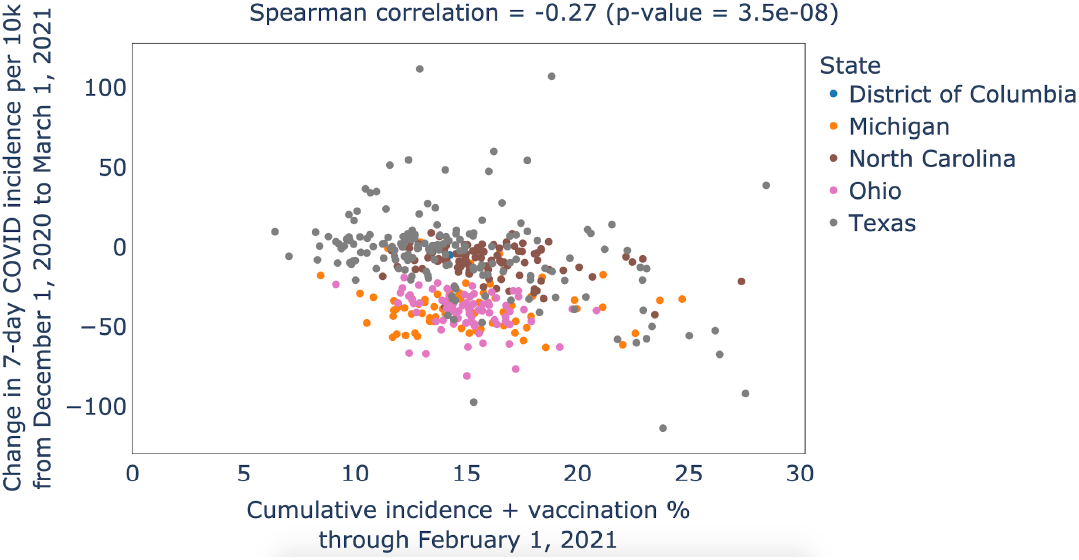
Cumulative vaccination through February 1, 2021 versus the difference between the 7-day COVID-19 incidence (per 10,000 individuals) between December 1, 2020 and March 1, 2021.

**Supplementary Figure S5.**
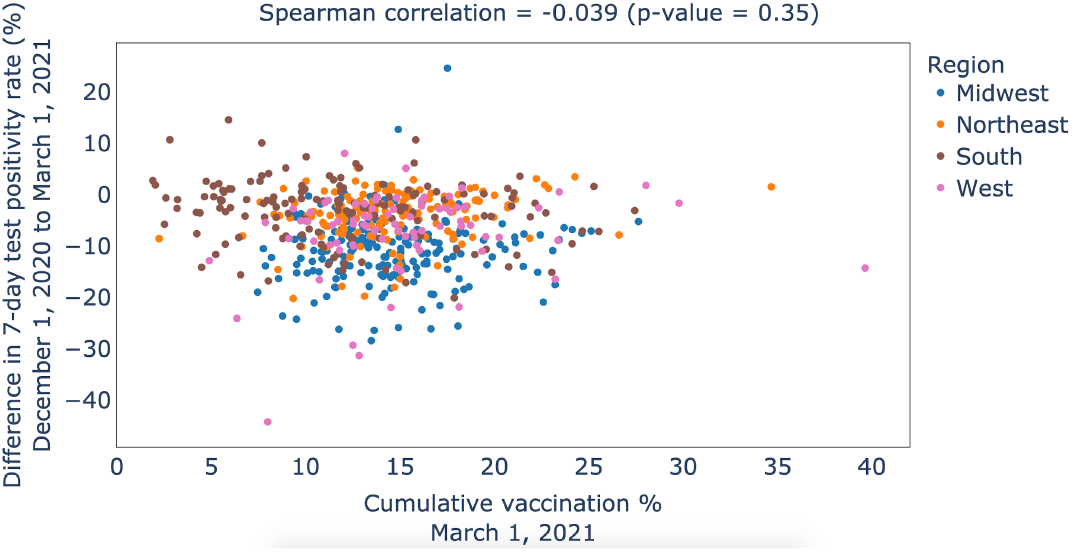
Cumulative county-level vaccination through March 1 2021 versus the change in 7-day percentage positivity rate (PPR) between December 1, 2020 and March 2021. This analysis includes 1987 counties from 40 states.

**Supplementary Figure S6.**
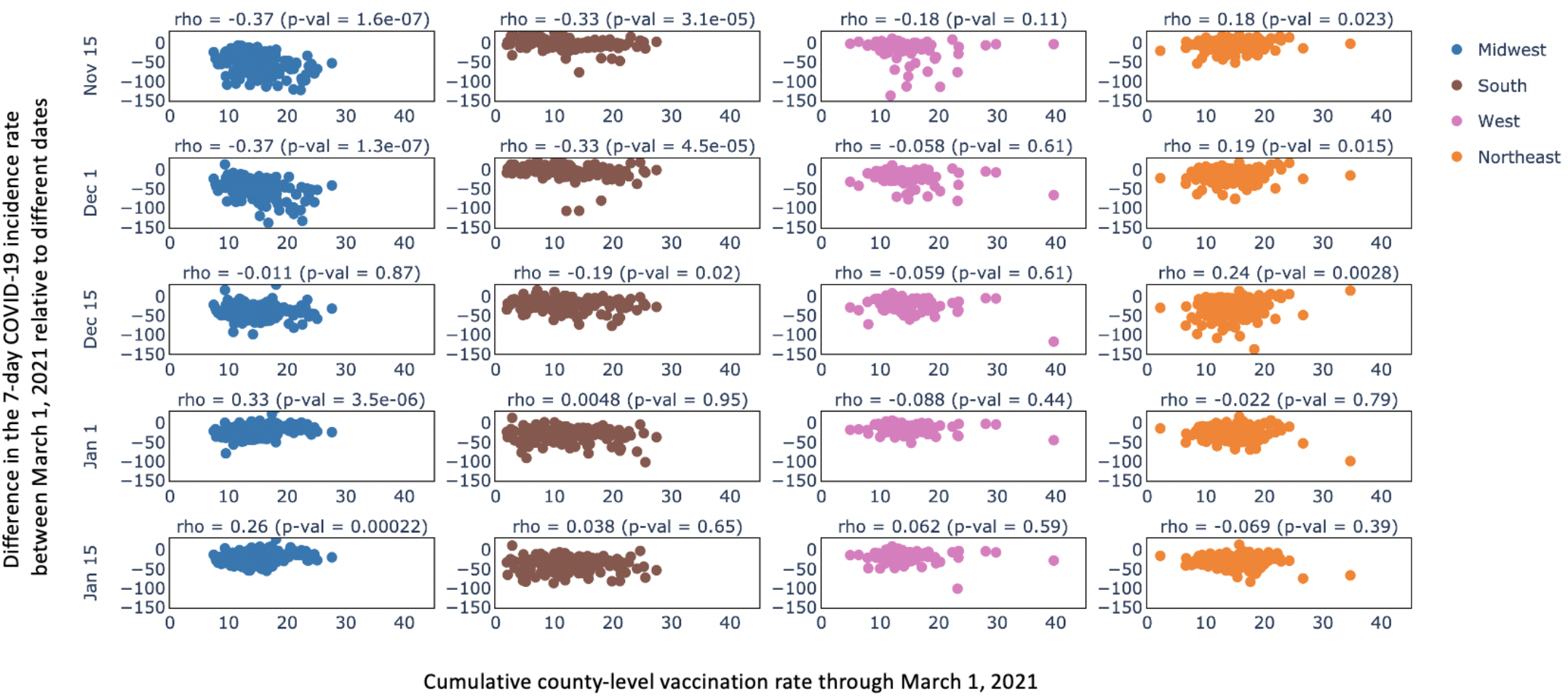
Matrix of scatter plots comparing cumulative county-level vaccination rate through March 1, 2021 with the Difference in the 7-day COVID-19 incidence rate between March 1, 2021 relative to difference dates (shown as rows). The columns correspond to the counties from different geographic regions: Midwest, South, West and Northeast.

## Footnotes

1 Dimmit county, Texas - https://public.nferx.com/covid-monitor-lab/vaccinationcheck?location=%5B48%2C48127%5D&page=1

2 Henry county, Ohio - https://public.nferx.com/covid-monitor-lab/vaccinationcheck?location=%5B39%2C39069%5D&page=1

3 Churchill county, Nevada - https://public.nferx.com/covid-monitor-lab/vaccinationcheck?location=%5B32%2C32001%5D&page=1

